# Do Sleep Quality Can Be the Intervening Factors of Personality Data to Occupational Fatigue?

**DOI:** 10.1101/2024.04.26.24306427

**Authors:** Doddy Izhar, David Kusmawan, Budi Aswin

## Abstract

Fatigue during work among oil and gas employees can have dangerous effects on wellbeing, economics, safety, and health. This cross-sectional survey was conducted in July and August of 2022 at two national oil and gas companies located in Muara Jambi and Jambi City. A convenience sample of 116 respondents was selected in total. To address the study hypotheses, partial least square-structural equation modeling (PLS-SEM) was employed. This study aims to determine the relationship between the risk of job weariness among Indonesian oil and gas workers in Jambi Province and the direct and indirect impacts of mental workload, sociodemographic characteristics, and sleep quality. Personality data has a significant and negative direct impact on occupational weariness at alpha 5% and with a path value of -0.203 (p-value: 0.047), corroborating the earlier hypothesis. For the second hypothesis, the path coefficient value of 0.462 (p-value: 0.000) clearly shows that sleep quality has an impact on occupational weariness. In order to improve sleep hygiene and address personality factors like age and length of employment, fatigue risk management strategies can be combined with those that are currently being used to control job tiredness.

## Introduction

Fatigue is a subject science studied in various research fields, such as cognitive neuroscience, exercise physiology, psychology, medical sciences, and public health (Skau *et al*., 2021). Fatigue is a non-specific symptom because it can be indicative of many causes or conditions including physiological states such as sleep deprivation or excessive muscular activity, medical conditions such as chronic inflammatory conditions, bacterial or viral infections, or autoimmune illnesses, psychiatric disorders such as major depression, anxiety disorders and somatoform disorders. Torres-Harding & Jason, 2005). In normal physiology, fatigue may be weakness or fatigue from repeated exertion or a decreased responses of cell, tissues, or organs after excessive stimulation, stress or activity (Hirshkowitz, 2013). Fatigue is the state of feeling very tired, weary or sleepy resulting from insufficient sleep, prolonged mental or physical work or extended periods of stress or anxiety, and repetitive task (Canadian Centre for Occupational Health and Safety, 2017). The feeling of fatigue is quite individual. Numerous interrelated aspects are evidently indicated by research. These have extremely diverse effects on various people. Other elements vary, such as the desire to work or avoid working overtime.

Fatigue has come to encompass many concepts. No single accepted definition. Multidimensional in nature such as acute or chronic, whole body or muscle level, physical or mental, central or peripheral nervous system, includes a decline in objective performance (Cavuoto & Megahed, 2016). Fatigue is a multidimensional construct that is associated with a range of physical, psychological, mental, and workplace risk factors. The effect of fatigue can be catastrophic and seriously jeopardize in workplace safety and health. Fatigue is a state of impairment that can negatively impact on safety, productivity, quality, morale, compliance, profits, and more. The economics loss due to occupational fatigue in US have been investigated (Ricci *et al*., 2007). Workers with fatigue cost employers $136.4 billion annually in health-related Lost Productive Time, an excess of $101.0 billion compared with workers without fatigue. Numerous well-publicized incidents, including Chernobyl, Exxon Valdes, and the Challenger Space Shuttle, have highlighted the role that fatigue plays. Furthermore, the oil and gas sector are inherently risky, and a number of widely reported investigations into significant incidents, including Piper Alpha (UK), Alexander Keilland (Norway), and Longford (Australia), have drawn the attention of management from both operating and contracting companies to safety concerns (Mearns & Yule, 2009).

Additionally, there is a correlation between worker health, wellbeing, and performance and fatigue on a variety of dimensions, including burnout, cognitive exhaustion related to tasks and sleep, physical fatigue, emotional fatigue, and perceptual fatigue (Yung *et al*., 2020). Furthermore, a number of studies have shown that fatigue may have a negative impact on the risk of cancer, mental health, and metabolic and cardiovascular health (Alroomi & Mohamed, 2021). It may also have an adverse effect on mood swings, job stress, cognitive decline, sleep disturbance, the incidence of MSDs (Heidarimoghadam *et al*., 2019), absenteeism from work (Sagherian et al., 2019), behavior, performance, and climate (Gu & Guo, 2022). Additionally, it may have an impact on reduced human performance levels, organizational productivity, and performance ability (Dahlan & Widanarko, 2022). Hazardous conduct leading to fatalities among oil and gas employees, likelihood of an incident, quality of life, wellbeing, and accident (Hinze *et al*., 2021). According to Benson et al. (2021), besides exhaustion, the oil and gas sector poses risks related to ergonomics (30%), physical hazards (26%), chemicals (23%), psychological hazards (18%), and biology (3%). Together with these other negative effects, fatigue also raised incident rates, medical expenses, employee turnover, forgetfulness, and a decline in alertness, reaction time, capacity for difficult job, and stress tolerance.

The result of the contemporary industrial civilization is fatigue (Caldwell, 2019). Workplace fatigue has emerged as a major problem, the leading cause of accidents and difficulties for Indonesian oil and gas employees. Primary risk factors clusters such as staff selection and training, monitoring and management, personal fatigue management, work pattern, staffing levels, shift work, circadian rhythm disruption, ultradian processes, health factors, mental and physical workload, sleep hygiene, sleep homeostasis, isolated work location, loneliness, lack of family support, education, BMI, long working duration, roster pattern, length of shift, timing of shift, insufficient recovery time between shifts, type of work, and insufficient rest breaks are some of the factors that contribute to occupational fatigue in the oil and gas and petrochemical industries. Extraneous variables such journey duration, social life, family obligations, and the number of children (Kang *et al*., 2021), (Rini *et al*., 2022), (Ghasemi *et al*., 2019), (Sinagabariang & Kurniawidjaja, 2024), (Garrubba & Joseph, 2019).

There are currently few association studies and mechanisms explaining how sociodemographic factors including age, length of employment, Body Mass Index (BMI), sleep quality, and mental workload affect job tiredness in oil and gas personnel. Additionally, by identifying the most important variables, fatigue risk management techniques should be used to mitigate and lower short- and long-term health and safety risks in oil and gas workers. The purpose of this study is to ascertain the association between the risk of work fatigue in oil and gas workers in Jambi Province, Indonesia, and the direct and indirect effects of sociodemographic factors, mental workload, and sleep quality.

## Method

### Research method

#### Study Design and Respondents

Two national oil and gas firms in Muara Jambi and Jambi City, Jambi Province, Indonesia, were the sites of this cross-sectional study, which was carried out in July and August of 2022. Convenience sampling was used to pick 116 respondents overall, all of whom were willing to work in the oil and gas industry. The Research Ethics Commission Faculty of Public Health UNAND University accepted this study (Ethic Number:19/UN16.12/KEP-FKM/2022). Respondents signed an informed consent form before to data collection, and their identity and confidentiality were ensured.

### Occupational Fatigue

The KAUPK1 questionnaire, consisting of 17 questions related to fatigue complaints, was prepared to evaluate the feelings of occupational fatigue in chronically fatigued Indonesian workers.

### Sleep Quality

The Pittsburgh Sleep Quality Index (PSQI) questionnaire contained 9 items, with 5 items consisting of 10 sub-items and 7 subscales including subjective sleep quality, sleep duration, sleep latency, sleep disturbance, habitual sleep efficiency, daytime dysfunction, and use of sleeping medication. Each dimension was weighted equally on a scale from 0 to 3 and the scores for all 7 dimensions were summed to produce a total score between 0 and 21, where a higher score indicated worse sleep quality.

### Mental Workload

The NASA-TLX questionnaire was developed by NASA Research Center (Hart & Staveland,1988) to measure the total mental workload of oil and gas workers. The NASA-TLX consisted of 6 items scale, which was used to evaluate different aspects of workload, Mental Demand (MD), Physical Demand (PD), Temporal Demand (TD), Effort (EF), Performance (PE), and Frustration Level (FR). There were two steps involved in measuring the workload. To ascertain which NASA-TLX dimensions were more important for carrying out their everyday tasks, respondents conducted a total of 15 pairwise comparisons across all 6 instrument dimensions in the first stage. The overall workload score would be calculated as the weighted average of the values for each dimension. This tool, which measured mental workload, was reliable and extensively utilized (Lund *et al*., 2021), (Tubbs-Cooley *et al*., 2018), (Mouzé-Amady *et al*., 2013).

### Data Analysis

To summarize the demographic data, descriptive statistics were employed. Partial Least Square-Structural Equation Modeling (PLS-SEM) was the method used to address the study hypotheses. On the recommendation of Götz *et al*. (2009), partial least squares (PLS-SEM) are taken into consideration. In their view, PLS analysis is a reliable measurement technique that can be used with small sample sizes and does not necessitate assumptions about the data’s measurement scale. This study intends to ascertain the effects of mental workload, sleep quality, and sociodemographic data (SD) on occupational fatigue (OF) with a particular focus on oil and gas employees.

In order to estimate complex models with several indicators and structural routes without making distribution assumptions about the data, the SEM-PLS method is highly appealing to a large number of researchers (Sarstedt *et al*., 2021).Furthermore applicable to smaller sample sets in which case the coefficient values according to Hair Jr *et al*. (2021), at the 5% significance level, 69 samples provide the minimal path likely to be significant, which is between 0.21 and 0.3.116 workers in Indonesia’s oil and gas industrial sector, specifically in the upstream segment, provided replies for the study.

The data analysis process employs selected statistical approaches, and the questionnaire follows a Likert scale format. The process of measuring the reflective model was called outer model analysis, and it involved multiple steps. These comprised evaluating the composite reliability, convergent and discriminant validity, and indicator loadings. Item loadings were restricted to values greater than 0.6. Higher than 0.6 Cronbach’s alpha values were seen as suggestive of a reliable internal consistency. Acceptable convergent validity was also shown by AVE values ≥ 0.05, while HTMT values < 0.85 were used to determine the results of the discriminant validity test.

Predictive Relevance (Q2) and the coefficient of determination (R2) were two of the metrics used to evaluate the inner model analysis. R2 values more than zero were significant, while values of 0.75, 0.50, and 0.25 were regarded as substantial, moderate, and weak, respectively. The value of probability from the bootstrap t-test was analyzed in order to determine the direct relationship between mental workload and sleep quality and occupational tiredness (Hair *et al*., 2019).

### Conceptual framework and hypothesis

**Figure 1.**
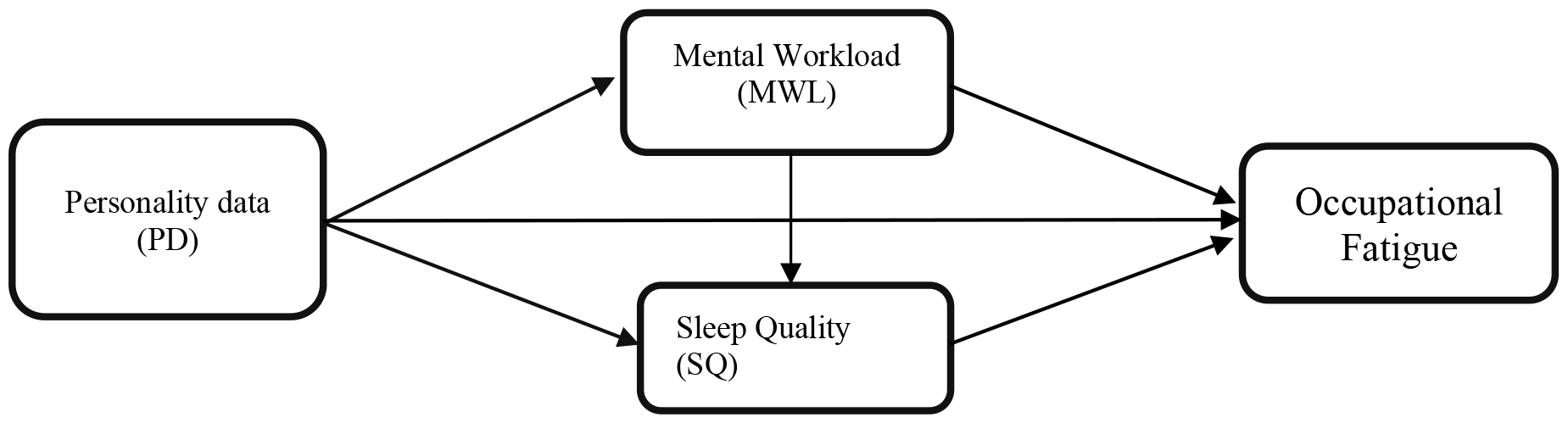
SEM conceptual framework.

## Result and Discussion

The distribution of respondents in Table 2. indicates that the percentage of job experience falls between 27 and 30 percent in all categories. The age group of 45 to 59 years old accounts for the largest amounts of workers (51.7%, or 60 individuals), followed by the 25–44 years age group, which has 50 individuals (43.1%). Ninety-three out of the 116 respondents, or 80.2% of the total, had an ideal (normal) BMI.

**Table 1.** Research Hypothesis

| Hypothesis | Relationship Description |
| --- | --- |
| H.1 | Personality data has a direct effect on occupational fatigue |
| H.2 | Sleep quality is an intervening factor of the effect of personality data to occupational fatigue |
| H.3 | Mental workload is an intervening factor of the effect of personality data to occupational fatigue |

**Table 2.** Distribution of PT X (Persero) Oil and Gas Workers Characteristics.

| Characteristics | Category | n | % |
| --- | --- | --- | --- |
| Working Periods/WP (year) | ≤ 9.5 | 29 | 25.0 |
|  | 9.25 - 17 | 30 | 25.9 |
|  | 17.01 - 24 | 30 | 25.9 |
|  | > 24 | 27 | 23.3 |
| Age (year) | < 15 | 0 | 0 |
|  | 15 - 24 | 6 | 5.2 |
|  | 25 - 44 | 50 | 43.1 |
|  | 45 - 59 | 60 | 51.7 |
| Body Mass Index | < 18.5 | 0 | 0 |
|  | 18.5 - 24.9 | 93 | 80.2 |
|  | 25 - 29.9 | 12 | 10.3 |
|  | ≥ 30 | 11 | 9.5 |
| Occupational Fatigue | Low | 52 | 44.8 |
|  | Hight | 64 | 55.2 |

### Evaluation of Model

Table 3 presents the findings, which indicate that the overall model goodness criteria in both the reflective measurement model and the structural model have been satisfied. In terms of dependability, each indicator’s loading factor value falls between 0.641 and 0.641. When the Heterotrait–Monotrait correlation value in Table 4 is less than 0.9 and the overall AVE value is greater than 0.5, the model can also be considered valid.

**Table 3.** Reflective Measurement and Structural Model.

| Latent Variable | Indicator | Reliability |  | Validity | VIF |  | R2 | R2-adj |
| --- | --- | --- | --- | --- | --- | --- | --- | --- |
|  |  | Outer loading | Cronbach's alpha | AVE | Outer | Inner |  |  |
| Personality Data | WP | 0.947 | 0.779 | 0.811 | 1.686 | 1.014 | 0.244 | 0.224 |
|  | Age | 0.852 |  |  | 1.686 |  |  |  |
| Mental Workload | N13 | 1.00 | 1.00 | 1.00 | 1.000' | 1.015 |  |  |
| Sleep Quality | P1 | 0.807 | 0.383 | 0.618 | 1.059 | 1.012 |  |  |
|  | P8 | 0.765 |  |  | 1.059 |  |  |  |
| Occupational Fatigue | K6 | 0.72 | 0.837 | 0.505 | 1.791 | - |  |  |
|  | K7 | 0.641 |  |  | 1.454 |  |  |  |
|  | K12 | 0.703 |  |  | 1.594 |  |  |  |
|  | K13 | 0.762 |  |  | 1.822 |  |  |  |
|  | K15 | 0.722 |  |  | 1.659 |  |  |  |
|  | K16 | 0.736 |  |  | 1.870 |  |  |  |
|  | K17 | 0.686 |  |  | 1.481 |  |  |  |

**Table 4.** Discriminant Validity.

| Variables | Mental Workload | Occupational fatigue | PD | Sleep Quality |
| --- | --- | --- | --- | --- |
| Mental Workload |  |  |  |  |
| Occupational Fatigue | 0.134 |  |  |  |
| Personality Data | 0.091 | 0.242 |  |  |
| Sleep Quality | 0.143 | 0.787 | 0.191 |  |

**Figure 2.**
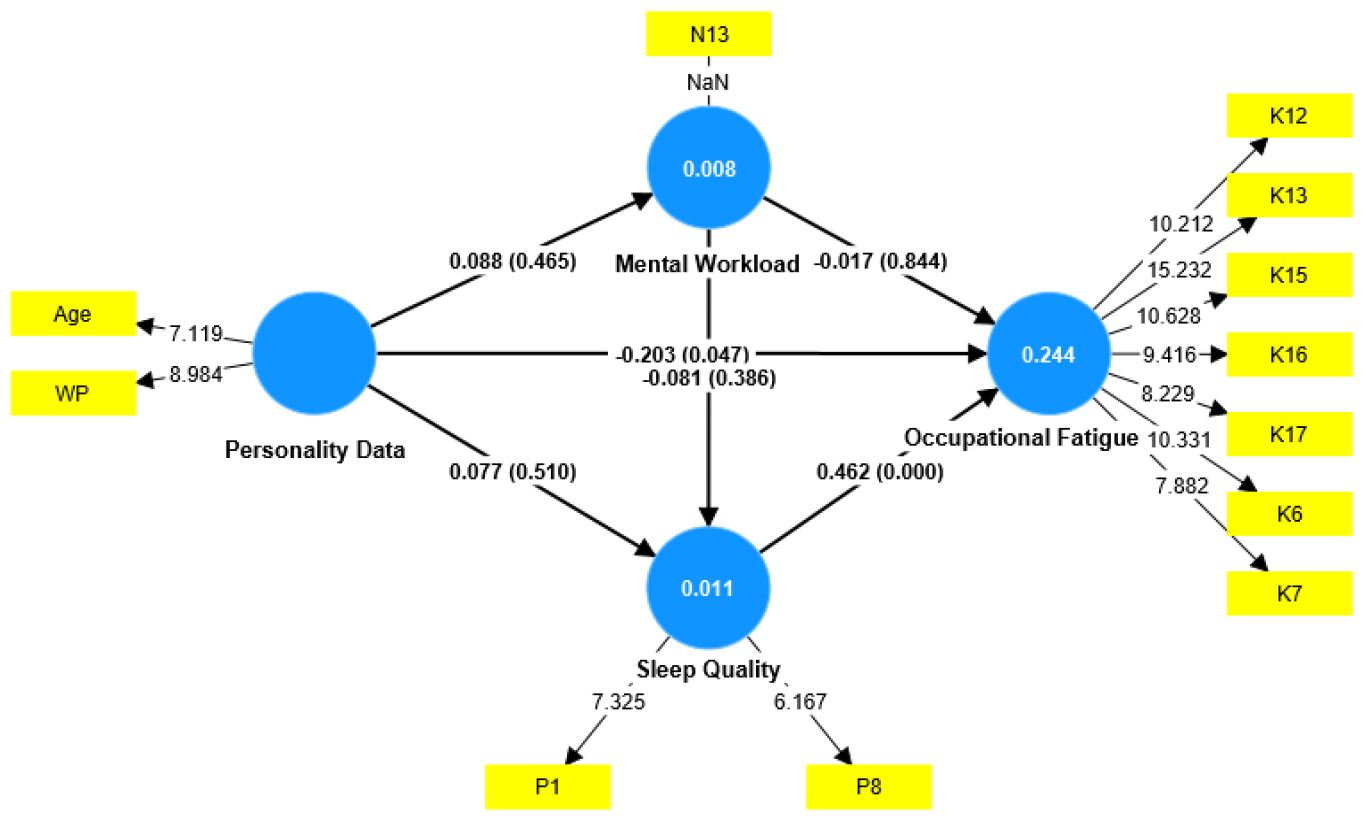
Conceptual Model SEM-PLS.

Table 5 provides evidence that, at alpha 5% and with a path value of -0.203 (p-value: 0.047), personality data directly influences occupational fatigue in a substantial and negative manner, supporting the first hypothesis. Similarly, the path coefficient value of 0.462 (p-value: 0.000) for the second hypothesis indicates that occupational fatigue is equivocally influenced by sleep quality.

**Table 5.** Hypothesis Test Results.

| Path |  | Coeff | T-stat | P-value |
| --- | --- | --- | --- | --- |
| From | To |  |  |  |
| Personality Data | Mental Workload | 0.088 | 0.730 | 0.465 |
| Personality Data | Sleep Quality | 0.077 | 0.659 | 0.510 |
| **Personality Data** | **Occupational Fatigue** | **-0.203** | **1.990** | **0.047\*** |
| Mental Workload | Sleep Quality | -0.081 | 0.866 | 0.386 |
| Mental Workload | Occupational Fatigue | -0.017 | 0.196 | 0.844 |
| **Sleep Quality** | **Occupational Fatigue** | **0.462** | **5.763** | **0.000\*** |

In this study, the third hypothesis was not proven, namely mental workload as a mediating factor in the relationship between the influence of personality data on work fatigue, which means that the occurrence of work fatigue in oil and gas workers in Jambi Province occurs directly due to personal data factors, namely length of service and age. This research is different from. In the latent variables of personality data, indicators of age and length of service have a negative influence on work fatigue. This means that work experience and age are inversely related to work fatigue. The longer the working period, the lower the influence on work fatigue among oil and gas workers in Jambi Province. In this study, productive age workers between 25 until 44 years age range (43.1%) felt greater work fatigue than those in the 45 to 59 years age range. Numerous research shows a confusing and contradictory link between occupational fatigue and aging. However, a few of this study have demonstrated a strong positive correlation between age and occupational fatigue. One intriguing finding of this research is that, as one ages, work-related weariness decreases and vice versa. It’s probable that workers older than 59 years old have greater abilities and adaption along with a somewhat lower mental and physical workload than this group of workers.

Age is one of the factors that affect an individual’s work ability. The hourly energy consumption on the condition of working muscles for each person is different, one of which is the age factor. Someone’s age factor will affect the basal metabolism will decrease and the individual will easily experience fatigue. Growing older was linked to worse recovery and higher levels of both acute and chronic fatigue. Every aspect of work-related exhaustion is impacted by aging, including changes in muscle strength (Giuliani *et al*., 2020).

Overall, muscle mass is determined by the equilibrium between the synthesis and breakdown of proteins, and both processes are influenced by a variety of variables, including hormonal balance, diet, physical activity, and illness (Frontera & Ochala,2015). Distinctive changes in the neuromuscular system and muscle action patterns (e.g., eccentric, concentric, and isometric) could impact fatigue recovery responses differently. Thompson *et al*., 2015). A significant decline in maximal voluntary isometric contraction occurs with aging due to neural activation of agonist muscles during knee extensors (Wu *et al*., 2016). It is commonly accepted that the muscles of the elderly contract with less force, relax more slowly, and show a downward shift in force-velocity correlations. Contractile velocity has a major impact on skeletal muscle’s age-related fatigue resistance (Callahan & Kent-Braun, 2011).

There is less of a direct impact on work tiredness the longer work periods. The work period is the amount of time an employee dedicates his strength to a certain organization, where the workforce can produce work that meets expectations based on aptitude, skills, and specific talents needed to complete the task at hand. Workers who have worked for a long time will typically feel more at ease in the firm since they have had enough time to acclimate to their surroundings and get along with other employees. Fatigue is a very personal sensation. Numerous interrelated aspects are evidently indicated by research. These have extremely diverse effects on various people. Other elements vary, such as the desire to work or avoid working overtime. P1 and P8, which are question indicators that gauge respondents’ typical bedtime (P1) and an evaluation of their overall sleep quality indicators (P8), are indicators on the sleep quality variable (PSQI) that have a little favourable impact on work tiredness. There is a higher chance of experiencing occupational fatigue among employees who start sleeping later in the evening. Similarly, for indication (P8), the likelihood of occupational fatigue increases with a worker’s opinion of their sleep quality.

The greatest way to combat occupational fatigue is to get enough sleep (Mehta et al., 2019). This is consistent with current research, which states that an increased risk factor for fatigue results from reduced rest duration and poor sleep quality. Fatigue was significantly impacted by the quality of sleep at 95% confidence interval. One theory was that employees were less likely to feel exhausted when they slept well. It has been determined that maintaining proper sleep hygiene is crucial for increasing employee productivity.

Sleep improves a wide range of mental processes in the brain, including memory, learning, and rational decision-making. Sleep is a kind gift to our mental health; it resets the emotional circuits in our brains, enabling us to face the social and psychological obstacles of the next day with poise and calm. Sleep replenishes the immune system’s stockpiles downstairs in the body, aiding in the fight against cancer, averting infection, and fending off a host of ailments. Sleep adjusts the insulin and glucose levels in the bloodstream to restore the body’s metabolic equilibrium. Our appetite is further controlled by sleep, which promotes healthy eating habits rather than hasty impulses when it comes to weight control. A healthy gut microbiota, which is where we know so much about our nutritional health starts, is maintained by getting enough sleep (Walker, 2017).

Due to its cross-sectional design, this study has some drawbacks common to this kind of research. One reason is that it is challenging to prove causation since, of course, long-term and experimental study is required to draw a definitive judgment about it. (Allison et al., 2022).

## Conclusion

In conclusion, the quality of sleep has a mediating role in the association between personality traits and occupational tiredness. There’s no denying that occupational fatigue is influenced by the quality of sleep. There is little doubt that the quality of sleep affects occupational fatigue. It is possible to combine fatigue risk management strategies with those currently in use to control work weariness in a way that enhances sleep hygiene and addresses personality factors like age and term of employment.

## Data Availability

All data produced in the present study are available upon reasonable request to the authors

## Acknowledgments

The support (project number 247/UN21.11/PT.01.05/SPK/2022) from the Directorate of Research and Community Engagement at Universitas Jambi is gratefully acknowledged by the authors.

## Notes

### Competing Interest Statement

The authors have declared no competing interest.

### Author Declarations

The Research Ethics Commission Faculty of Public Health UNAND University accepted this study (Ethic Number:19/UN16.12/KEP-FKM/2022).

